# PCR Detection and Species Identification of Leishmania in Microscopy-Negative Smears from Suspected Cutaneous Leishmaniasis in Iran

**DOI:** 10.64898/2026.09.17.26363305

**Authors:** Fatemeh Yadollahzadeh Chari, Mehdi Fakhar

**Affiliations:** Department of Medicine, Mazandaran University of Medical Sciences, Sari, Iran; Department of Parasitology and Medical Mycology, Mazandaran University of Medical Sciences, Sari, Iran

**Author notes:** Corresponding author: Fatemeh Yadollahzadeh Chari, Mazandaran University of Medical Sciences, Sari, Iran.

**Keywords:** cutaneous leishmaniasis, *Leishmania major*, *Leishmania tropica*, PCR, kDNA, microscopy-negative smear, Iran

## Abstract

**Background:** Direct microscopy remains a practical first-line test for cutaneous leishmaniasis (CL), but it can miss infections when parasite numbers are low or lesions are chronic. We evaluated whether PCR could detect and identify *Leishmania* in Giemsa-stained smears that had already been reported as microscopy-negative.

**Methods:** We retrospectively analyzed 131 microscopy-negative smears from patients with clinically suspected CL who attended health-center laboratories in Gonbad-e-Qabus and Torbat-e-Jam, Iran, between March 2013 and February 2015. DNA was recovered from the stained slides by phenol-chloroform-isoamyl alcohol extraction. kDNA PCR was used for molecular detection and species identification.

**Results:** *Leishmania* DNA was detected in 34 of 131 microscopy-negative samples (25.9%; exact 95% CI, 18.7%-34.3%). Twenty-three positive samples (67.6%) were identified as *L. major* and 11 (32.4%) as *L. tropica*. All *L. major*-positive samples came from Gonbad-e-Qabus, whereas all *L. tropica*-positive samples came from Torbat-e-Jam. Most PCR-positive lesions (27/34, 79.4%) were chronic, secondarily infected, and/or clinically atypical. In a small laboratory verification panel, all 10 microscopy-positive smears were PCR-positive and all 10 non-leishmanial dermatosis controls were PCR-negative.

**Conclusions:** About one in four clinically suspected cases with a negative smear still had detectable *Leishmania* DNA. PCR of previously stained slides can therefore provide useful second-line confirmation and species information, especially when lesions are chronic or atypical. The small control panel supports assay performance in the study laboratory but is not large enough to provide precise estimates of clinical sensitivity or specificity.

## 1. Introduction

Cutaneous leishmaniasis (CL) continues to place a substantial burden on endemic communities. WHO surveillance for 2024 recorded 211,466 new CL cases worldwide, with most reported cases concentrated in the Eastern Mediterranean Region and the Americas [1]. As of July 2026, Iran remained among the countries reporting more than 5,000 CL cases to WHO [2]. These figures are a reminder that CL is not only a historical tropical disease; it remains a current diagnostic and public-health problem in Iran and neighboring countries.

The clinical diagnosis can be deceptively difficult. CL may resemble bacterial or fungal infection, cutaneous tuberculosis, inflammatory dermatoses, or even neoplastic disease. Direct examination of Giemsa-stained lesion material is inexpensive and highly specific when amastigotes are seen, but a negative smear does not reliably exclude infection. Sampling quality, lesion age, low parasite burden, secondary infection, and examiner experience all affect microscopy yield. Recent reviews continue to identify these limitations as an important cause of missed or delayed diagnosis [3,4].

Molecular tests can help close that gap. PCR can detect small amounts of parasite DNA, and the choice of target can also allow species identification. Recent reviews and meta-analyses report strong overall performance for PCR-based diagnosis, while also highlighting substantial variation in extraction methods, molecular targets, platforms, and validation standards [4,5]. kDNA minicircles are particularly attractive because they occur in high copy number. WHO has also moved toward harmonizing PCR procedures and quality assurance for skin-related neglected tropical diseases [6].

Species identification adds information that microscopy cannot provide. In Iran, *L. major* and *L. tropica* account for most CL, but their geographic distribution is uneven [7–9]. This distinction is relevant for epidemiology and can matter clinically because species differ in natural history, treatment response, and the likelihood of difficult-to-treat disease [3]. A related study from the same research program showed that DNA could be recovered from stained slides and used for species-specific PCR in patients from Pakdasht, Iran [10].

We therefore asked a simple, clinically relevant question: among patients whose lesions looked suspicious for CL but whose direct smears were negative, how often could *Leishmania* still be detected from the original stained slide, and which species were present? Because the samples were collected in 2013-2015, our aim is not to describe current regional prevalence. Instead, we focus on the diagnostic value of archived stained smears and the additional yield of PCR after negative microscopy.

## 2. Materials and Methods

### 2.1 Study design and setting

This retrospective laboratory study used archived Giemsa-stained smears from patients with skin lesions clinically suspected to represent CL who attended the health-center laboratories of Gonbad-e-Qabus (Golestan Province) and Torbat-e-Jam (Razavi Khorasan Province), Iran, between March 2013 and February 2015. The source study recorded age, sex, residence, lesion onset, number and location of lesions, occupation, travel or residence history, and lesion morphology. We included 131 smears that had been reported as negative for *Leishmania* amastigotes by direct microscopy. The original microscopy was performed by experienced health-center personnel, and the slides were re-examined by light microscopy before molecular testing.

### 2.2 DNA extraction from stained smears

DNA was recovered directly from the stained material on each slide. Approximately 200 µL of lysis buffer was spread over the smear, and the loosened material was transferred to a 2-mL microtube. Proteinase K (15 µL per 200 µL of lysate; 20 mg/mL) was added, followed by incubation at 56°C for 2 h or at 37°C overnight. An equal volume of phenol-chloroform-isoamyl alcohol was then added. After centrifugation at approximately 13,000-15,000 rpm for 10-15 min, the aqueous phase was transferred to a clean tube. DNA was precipitated with cold absolute ethanol at −20°C, washed, dried, and resuspended in 50 µL of sterile distilled water. DNA quality was checked by agarose gel electrophoresis and/or spectrophotometry according to the original laboratory protocol.

### 2.3 kDNA PCR and species identification

Two conventional PCR assays targeting *Leishmania* kDNA minicircle sequences were used. For molecular detection, RV1 (5′-CTT TTC TGG TCC CGC GGG TAG G-3′) and RV2 (5′-CCA CCT GGC CTA TTT TAC ACC A-3′) were used, with an expected product of approximately 145 bp in the study protocol. Each 25-µL reaction contained 12.5 µL of commercial master mix, 1 µL of each primer, 5.5 µL of sterile distilled water, and 5 µL of template DNA. Cycling conditions were 94°C for 4 min; 35 cycles of 94°C for 40 s, 57°C for 40 s, and 72°C for 30 s; followed by 72°C for 3 min.

Species identification used the LINR4/LIN17 primer pair targeting variable kDNA minicircle sequences. The primer sequences are reported here according to the published assay used by this laboratory program: LINR4, 5′-GGG GTT GGT GTA AAA TAG GG-3′; LIN17, 5′-TTT GAA CGG GAT TTC TG-3′ [14,16]. The source thesis contains a transcription error in the LIN17 sequence, so the published assay sequence is used in this manuscript. Reference strains *L. tropica* MHOM/IR/89/ARD2 and *L. major* MRHO/IR/75/ER were included. Cycling conditions were 94°C for 4 min; 40 cycles of 94°C for 45 s, 52°C for 45 s, and 72°C for 50 s; followed by 72°C for 5 min. Products were separated on 1.5% agarose gel. Under the study conditions, *L. major* and *L. tropica* produced bands of approximately 650 bp and 760 bp, respectively.

### 2.4 Control samples

The original study included a small laboratory verification panel. Ten smears in which *Leishmania* amastigotes had been identified by microscopy served as positive controls. Ten smears from patients with non-leishmanial skin disease, including dermatophytosis and cutaneous tuberculosis, served as negative disease controls. These samples were used to confirm expected assay behavior and were not treated as a representative diagnostic-accuracy cohort.

### 2.5 Statistical analysis

The source database was analyzed in SPSS version 16.0 using chi-square tests. For this reconstructed manuscript, the primary analyses are descriptive because the complete patient-level dataset was not available from the thesis record. Exact binomial 95% confidence intervals (CIs) are reported for the main proportions. Counts and percentages were retained only when their denominators were internally consistent in the source thesis; variables with conflicting denominator information were not re-analyzed.

### 2.6 Ethical approval

The project was reviewed and approved by the Research Ethics Committee of Mazandaran University of Medical Sciences (IR.MAZUMS.REC.1395.34). The approval document corresponds to the project titled “Evaluation of PCR for the Detection of *Leishmania* Amastigotes in Negative Smears from Patients with Cutaneous *Leishmania*sis.” Only de-identified, aggregate information is reported in this manuscript.

## 3. Results

### 3.1 PCR detection among microscopy-negative cases

The 131 microscopy-negative suspected cases included 75 males (57.2%) and 56 females (42.8%), with ages ranging from 2 to 75 years. Seventy-one samples (54.1%) came from Gonbad-e-Qabus and 60 (45.9%) from Torbat-e-Jam. PCR detected *Leishmania* DNA in 34 of the 131 smears (25.9%; exact 95% CI, 18.7%-34.3%). The remaining 97 samples (74.1%) were PCR-negative. In practical terms, PCR identified evidence of infection in roughly one quarter of cases that had been negative by direct microscopy.

**Table 1.** PCR detection and species distribution by study site.

| Study site | Microscopy-negative samples, n | PCR-positive, n (%) | 95% CI | Species among PCR-positive samples |
| --- | --- | --- | --- | --- |
| Overall | 131 | 34 (25.9) | 18.7-34.3 | *L. major* 23; *L. tropica* 11 |
| Gonbad-e-Qabus | 71 | 23 (32.4) | 21.8-44.5 | *L. major* 23 |
| Torbat-e-Jam | 60 | 11 (18.3) | 9.5-30.4 | *L. tropica* 11 |

### 3.2 Species and clinical characteristics of PCR-positive patients

Among the 34 PCR-positive samples, 23 (67.6%; exact 95% CI, 49.5%-82.6%) were identified as *L. major* and 11 (32.4%; exact 95% CI, 17.4%-50.5%) as *L. tropica*. The site pattern was complete: all *L. major*-positive samples were from Gonbad-e-Qabus, whereas all *L. tropica*-positive samples were from Torbat-e-Jam.

Twenty-one PCR-positive patients (61.8%) were male and 13 (38.2%) were female. The largest age group was 15-30 years (13/34, 38.2%), and 19 patients (55.9%) had a single lesion. Lesions were most often on the hands (45.8%), followed by the feet (28.1%), face (10.4%), and other body sites (15.7%). Notably, 27 of the 34 PCR-positive lesions (79.4%) were described as chronic, secondarily infected, and/or atypical, while only 7 (20.6%) were acute and typical. Farmers were the largest single occupational group (12/34, 35.3%).

**Table 2.** Selected characteristics of the 34 PCR-positive cases.

| Characteristic | Category | n (%) |
| --- | --- | --- |
| Sex | Male | 21 (61.8) |
|  | Female | 13 (38.2) |
| Age group | <5 years | 2 (5.9) |
|  | 5-15 years | 8 (23.5) |
|  | 15-30 years | 13 (38.2) |
|  | 30-45 years | 5 (14.7) |
|  | >45 years | 6 (17.7) |
| Number of lesions | 1 | 19 (55.9) |
|  | 2 | 8 (23.5) |
|  | 3 | 5 (14.7) |
|  | >3 | 2 (5.9) |
| Lesion site | Hand | 45.8% |
|  | Foot | 28.1% |
|  | Face | 10.4% |
|  | Other | 15.7% |
| Lesion morphology | Acute/typical | 7 (20.6) |
|  | Chronic, secondarily infected and/or atypical | 27 (79.4) |

### 3.3 Representative species-specific PCR products

The species-specific assay produced bands that matched the reference strains used in the study: approximately 760 bp for *L. tropica* and 650 bp for *L. major*. A representative gel from the original thesis is shown in Figure 3.

**Figure 1.**
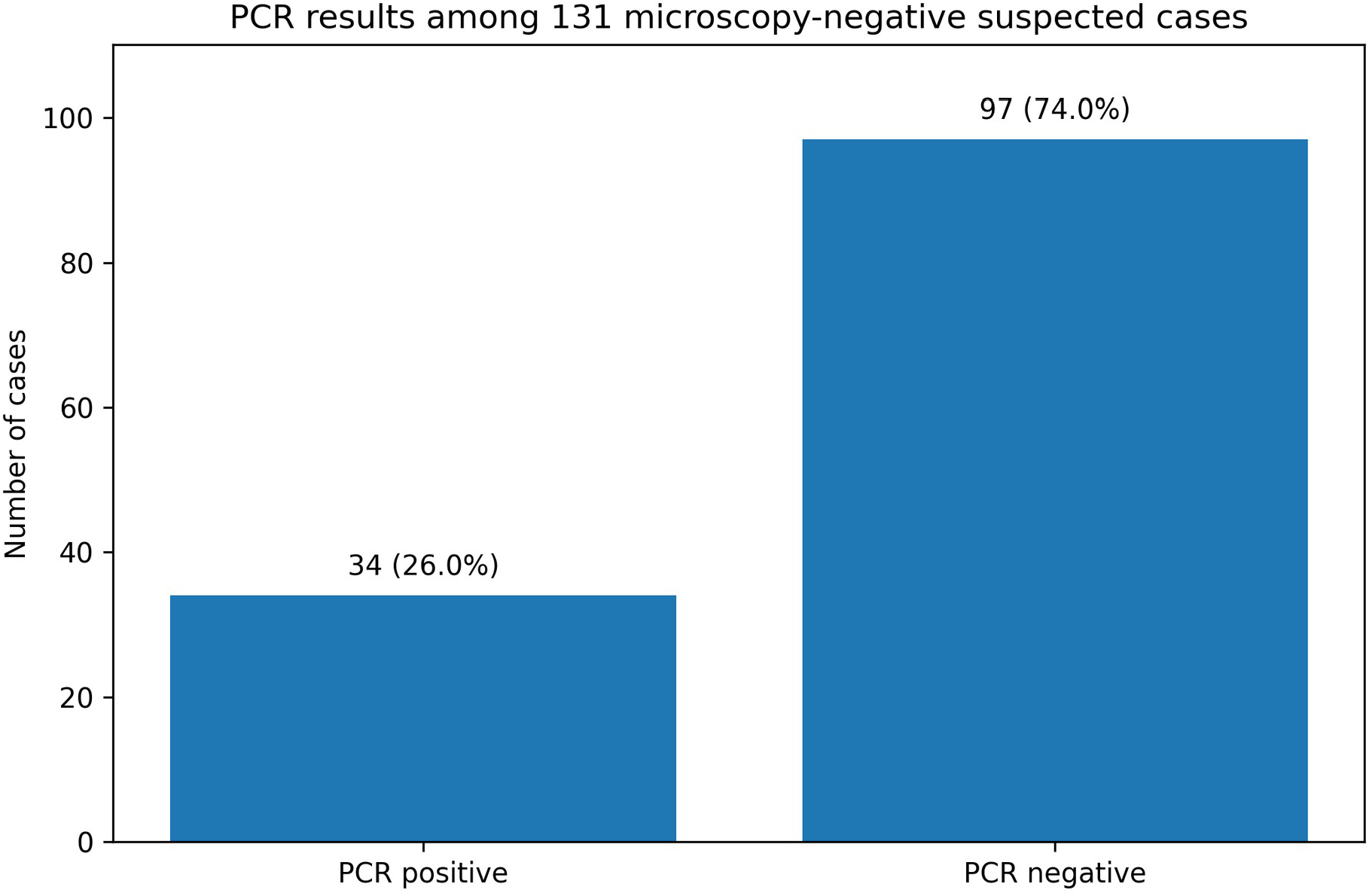
PCR results among 131 clinically suspected cutaneous leishmaniasis cases whose direct Giemsa-stained smears were negative by microscopy.

**Figure 2.**
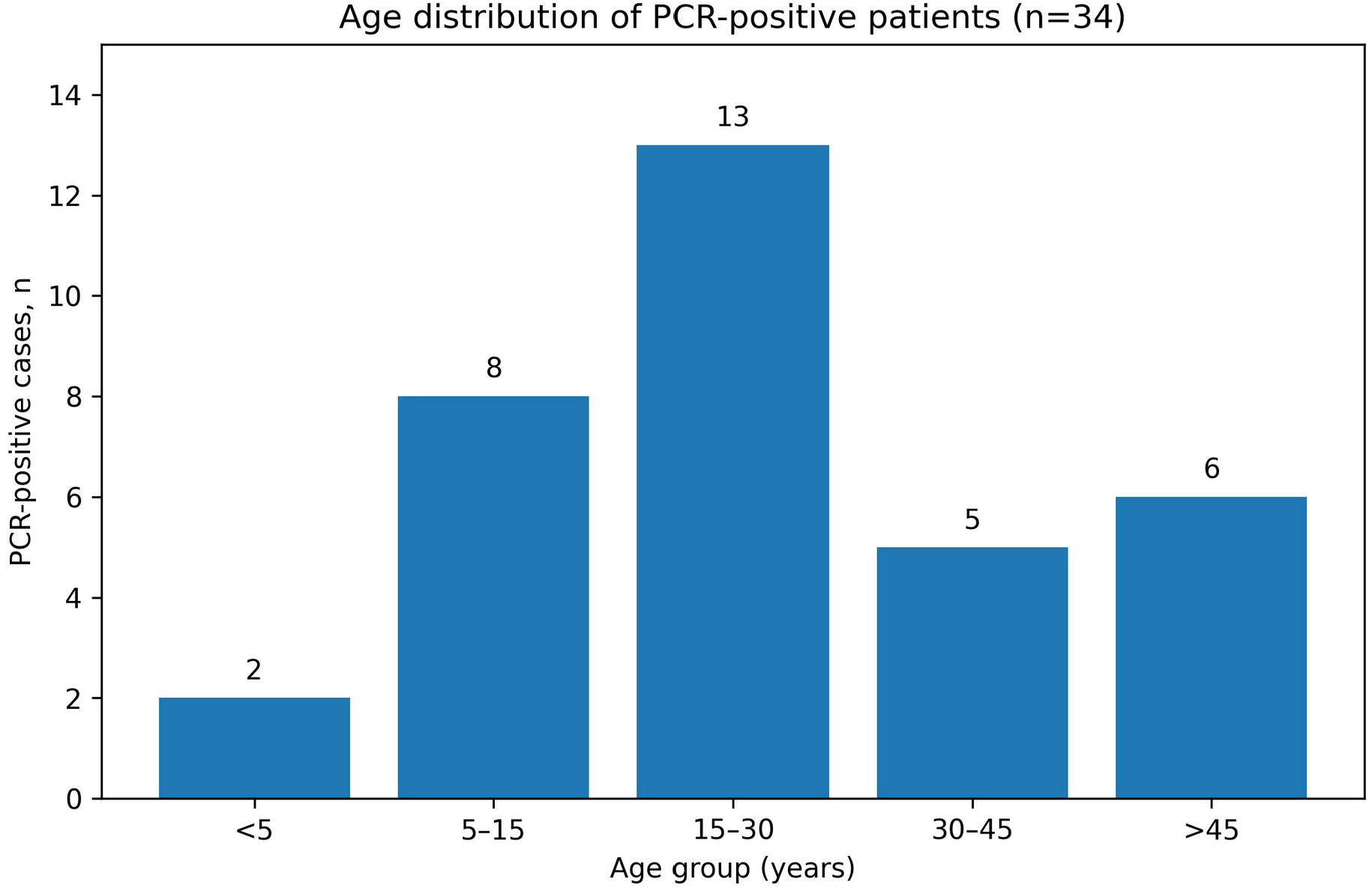
Age distribution of the 34 PCR-positive patients. Counts are reproduced from the source thesis.

**Figure 3.**
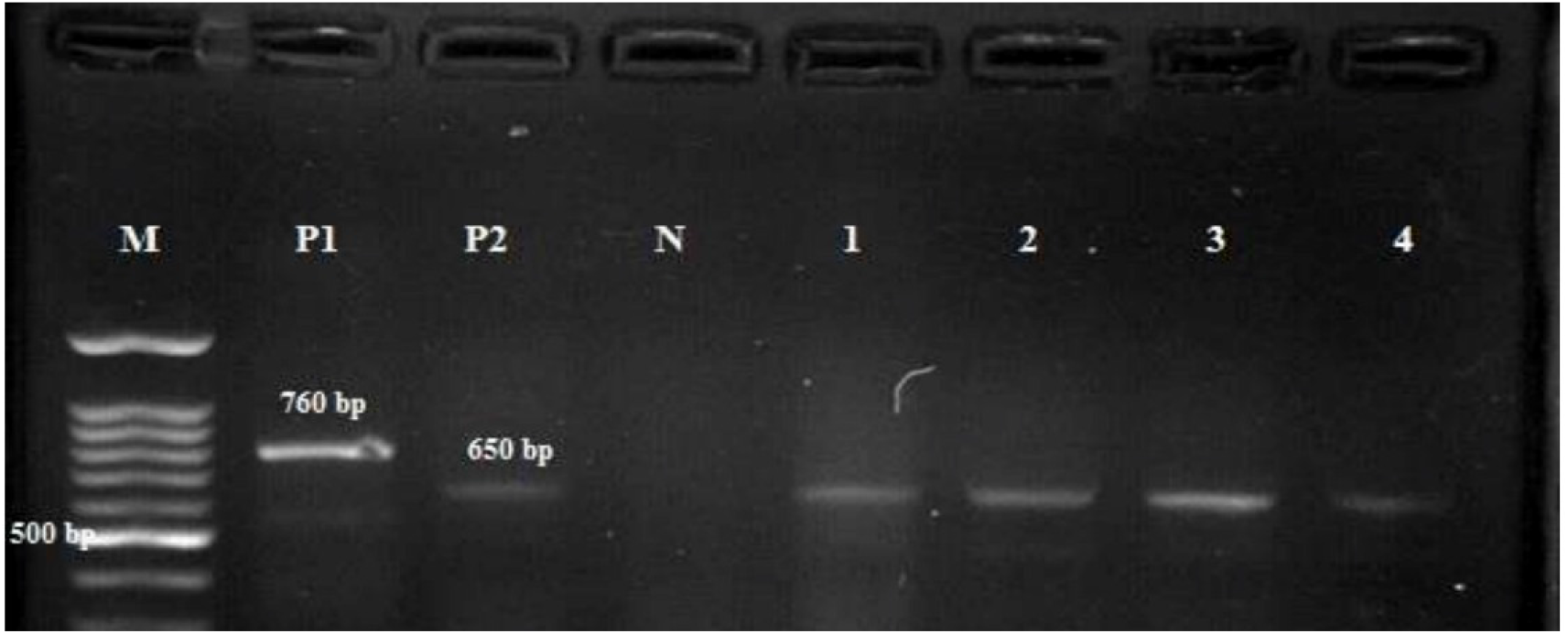
Representative agarose gel from the species-specific LINR4/LIN17 PCR. M, 100-bp DNA marker; P1, *L. tropica* reference strain (approximately 760 bp); P2, *L. major* reference strain (approximately 650 bp); N, negative control; lanes 1-4, representative patient samples. Image reproduced from the original MD thesis.

### 3.4 Laboratory verification panel

All 10 microscopy-positive control smears were PCR-positive, and all 10 non-leishmanial dermatosis controls were PCR-negative. Although each observed proportion was 100%, the exact 95% CI is wide (approximately 69.2%-100%) because only 10 samples were included in each group. We therefore interpret these findings as laboratory verification rather than as precise estimates of clinical sensitivity and specificity.

## 4. Discussion

PCR detected *Leishmania* DNA in 25.9% of patients whose lesions were clinically suspicious for CL but whose direct smears were negative. Put another way, one in four negative smears in this selected group still contained detectable parasite DNA. That is the clearest message of the study and the reason the findings remain useful despite the age of the dataset.

The result is consistent with earlier work on difficult or smear-negative samples. Mohaghegh et al. detected *Leishmania* DNA in 9 of 81 (11.1%) negative stained smears [11], Omidian et al. reported PCR positivity in 5 of 30 (16.7%) clinically suspected smear-negative lesions [12], and Fakhar et al. detected parasite DNA in 35 of 62 (56.4%) microscopy-negative smears in a referral population [13]. The range is wide, which is not surprising: lesion duration, sampling technique, referral patterns, extraction method, molecular target, and laboratory workflow all influence yield. Our 25.9% positivity rate falls comfortably within that broader experience.

The clinical profile of the PCR-positive patients also helps explain why microscopy failed. Nearly four fifths of the positive lesions were chronic, secondarily infected, and/or atypical. As lesions mature, parasite numbers often fall, while crusting, inflammation, bacterial or fungal superinfection, and difficult sampling make direct visualization even harder. PCR does not require an intact amastigote to be visible in the microscope field; it only requires amplifiable parasite DNA. Contemporary reviews and meta-analyses support this advantage, while also stressing that molecular performance depends on specimen type, extraction, target sequence, and assay standardization [3–5].

An additional strength of this approach is that it used the slide already collected for microscopy. Recovering DNA from a stained slide avoids a second scraping or biopsy and can still be useful when the patient has left the clinic. That practical feature is relevant to reference laboratories and retrospective investigations, and it fits well with current WHO efforts to harmonize PCR procedures and quality assurance for skin-related neglected tropical diseases [6].

The species results followed a clear geographic pattern. All PCR-positive samples from Gonbad-e-Qabus were *L. major*, whereas all positives from Torbat-e-Jam were *L. tropica*. This agrees with the broader molecular epidemiology of CL in Iran, where *L. major* and *L. tropica* predominate but their relative frequency changes substantially by region [7–9]. A national meta-analysis reported pooled frequencies of 67.3% for *L. major* and 32.1% for *L. tropica* among CL isolates [7], proportions strikingly similar to those observed in our PCR-positive group, although our study was not designed to estimate national or contemporary prevalence.

The related Pakdasht study from the same laboratory program provides useful methodological context. That study also recovered kDNA from stained slides using phenol-chloroform-isoamyl alcohol extraction and used species-specific PCR, identifying *L. major* in all characterized cases [10]. The present work applies a similar laboratory strategy to a more difficult diagnostic group: patients selected specifically because microscopy had been negative.

The small verification panel should not be overinterpreted. PCR was positive in all 10 microscopy-positive controls and negative in all 10 disease controls, but groups of 10 are far too small to support a robust claim of 100% clinical sensitivity or specificity. The main contribution of this study is therefore the additional diagnostic yield among smear-negative suspected cases, not a definitive head-to-head accuracy estimate.

### 4.1 Strengths and limitations

The study has several strengths. It focused on a clinically important problem, used the original stained slides rather than requiring repeat sampling, included two epidemiologically distinct study sites, and provided species-level identification. It also preserves a useful historical dataset from endemic settings and demonstrates that archived routine diagnostic material can support molecular testing.

The limitations are equally important. The main cohort included only microscopy-negative suspected cases, so the primary dataset cannot provide conventional sensitivity, specificity, positive predictive value, or negative predictive value for PCR versus microscopy. The positive and negative verification groups contained only 10 samples each. Species assignment was based on conventional endpoint PCR and expected band size rather than sequencing. Complete patient-level data were not available for this manuscript reconstruction, which limited multivariable analyses and prevented independent re-analysis of every chi-square comparison reported in the thesis. Finally, the samples were collected in 2013-2015, so the species pattern should not be interpreted as a current prevalence estimate for either city.

## 5. Conclusion

In this two-center study, kDNA PCR detected *Leishmania* DNA in 34 of 131 (25.9%) clinically suspected CL cases whose direct smears were negative. *L. major* was identified in Gonbad-e-Qabus and *L. tropica* in Torbat-e-Jam. These findings support PCR of Giemsa-stained slides as a practical second-line approach when microscopy is negative, particularly for chronic or atypical lesions, while also providing species information. Because the study used historical samples, its value lies in the diagnostic workflow and incremental yield rather than in describing current regional prevalence.

## Data Availability

The de-identified data underlying the findings of this study are available from the corresponding author upon reasonable request, subject to applicable institutional and ethical requirements.

## Declarations

### Ethics approval

The project was approved by the Research Ethics Committee of Mazandaran University of Medical Sciences (IR.MAZUMS.REC.1395.34). Only de-identified aggregate information is reported in this manuscript.

### Consent for publication

Not applicable; no identifiable patient information is presented.

### Data availability

The aggregate data supporting the findings are reported in this article and the source MD thesis. The original patient-level dataset is not included with the preprint. Archived study materials, where available, may be requested from the corresponding author subject to institutional and ethical requirements.

### Competing interests

The authors declare no competing interests.

### Funding

This work was conducted as part of an MD thesis at Mazandaran University of Medical Sciences. No external commercial funding was reported in the source thesis.

### Authors’ contributions

F.Y.C. carried out the thesis research, laboratory work, data collection, interpretation, and manuscript drafting. M.F. contributed to study conception, methodology, supervision, interpretation, and critical revision of the manuscript. Both authors reviewed and approved the final manuscript.

## Acknowledgements

The authors thank Hajar Ziaei Hezarjaribi for her advisory role in the original MD thesis and the staff of the health-center laboratories in Gonbad-e-Qabus and Torbat-e-Jam for their support. This manuscript was developed from the MD thesis of Fatemeh Yadollahzadeh Chari and updated for contemporary scientific reporting.

## Notes

### Competing Interest Statement

The authors have declared no competing interest.

### Author Declarations

The University/Regional Research Ethics Committee in Biomedical Research of Mazandaran University of Medical Sciences gave ethical approval for this work (approval number IR.MAZUMS.REC.1395.34).

